# Bridging health registry data acquisition and real-time data analytics

**DOI:** 10.1101/2024.06.12.24308496

**Authors:** Johannes Schmidt, Sita Arjune, Volker Böhm, Roman-Ulrich Müller, Philipp Antczak

**Affiliations:** Bonacci GmbH, Robert-Koch-Straße 8, 50937 Köln; Department II of Internal Medicine and Center for Molecular Medicine Cologne, University of Cologne, Faculty of Medicine and University Hospital Cologne, Cologne, Germany; Center for Rare Diseases Cologne, Faculty of Medicine and University Hospital Cologne, University of Cologne, Cologne, Germany; Cologne Excellence Cluster on Cellular Stress Responses in Aging-Associated Diseases (CECAD), Cologne, Germany; Institute for Genetics, University of Cologne, Cologne, Germany

## Abstract

The number of clinical studies and associated research has increased significantly in the last few years. Particularly in rare diseases, an increased effort has been made to integrate, analyse, and develop new knowledge to improve patient stratification and wellbeing. Clinical databases, including digital medical records, hold significant amount of information that can help understand the impact and progression of diseases. Combining and integrating this data however, has provided a challenge for data scientists due to the complex structures of digital medical records and the lack of site wide standardisation of data entry. To address these challenges we present a python backed tool, Meda, which aims to collect data from different sources and combines these in a unified database structure for near real-time monitoring of clinical data. Together with an R shiny interface we can provide a near complete platform for real-time analysis and visualization.

## Introduction

The medical world has seen a paradigm shift in recent years, acknowledging that data collection and analysis is key to understand the most pressing challenges in human health. Particularly with rare diseases, where the low number of patients impact the statistical analysis of these, must ensure that high quality and systematic collection of data is optimised. Often retrospectively collected data is available within the hospitals medical record systems but are plagued by numerous free-text fields, simple collection of laboratory values where the measurement units are not standardised across the fields, and the sheer amount of variables that that have been accumulated into these databases over years of use. Transitioning such database entities to more standardised and usable structures for clinical research or even simple oversight of departments within a healthcare organisation can prove to be challenging and associated with a very high cost of implementation and transition.

Furthermore, quality control of such data is often performed only when data is extracted for clinical research and entry failures only noticed when compared to other individuals. This proves one of the major headaches for data scientists who aim to integrate and analyse such data in various contexts. Within the medical field, and especially for laboratory values, thresholds are known that describe compatibility with life, giving a first indication whether the values entered are reasonable. Given the broad spectrum of diseases and health states in humans it is not reasonable to assume that each medical professional knows and applies these thresholds, particularly when they are early in their career. Written laboratory reports often include the range and thresholds to consider, but once provided within the database these are lost or stored in such a way that they are not directly accessible by the user. A more direct, and disease tailored, approach on the level of medical record oversight and data entry could lead to improved data quality and medical understanding.

In the last decade in Germany, there has been significant progress in the development of standardised interfaces to allow interoperability of data between health care institutions. The FHIR interface aims to provide a solution to transport data from one location to another and allow the sharing of patient data. While these developments are of great importance in the medical field, they do not fully address the internal and integrative use in clinical research. To this end, we have developed a small highly flexible and dynamic tool to collate, aggregate, integrate, and visualize clinical data. We opted to develop a centralized database structure, that pulls in data from multiple sources, formats, aligns, and tests them to ensure highest possible data quality. This database can then be connected to a visualization framework such as R shiny, Grafana, or Tableau to present the data in an aggregated fashion to healthcare professionals.

## Implementation

### Development of a universal translation service for medical data (MEDA)

Clinical registries are often based on data registration, management and storage designs which lack up-to-date database standards. These range from mere spreadsheets to specialised but non-standardised databases from various providers to collect and represent data. While these web-based tools often contain the ability to validate data entry, or limit the entry to specific datatypes, these features are often not used due to their complex configuration or lack of knowledge and experience by the initiating user. In addition, database structures are often inefficiently designed and variable names lack the descriptive nomenclature which allows other users to understand their values and implement these variables in their analyses. This then often requires the development of additional variable-dictionaries which provide extended definitions of the values. Particularly for the key aim of such datasets, i.e. downstream biostatistical analyses or real-time visualisation, the initial data structure and simplicity of the database is an important aspect for implementation and use. Live data visualization for both data sharing and in-house observation of cohort development is hardly possible in this setting. This is especially important when the developer of the database has left the organisation and the approaches and thoughts during the development process have not been documented accordingly. In most cases, the initial design allows questions posed by the developer and researchers associated with the project to be answered, however they can hinder the further use and analyses of these important data.

To address the challenges around clinical datasets described above, and to enable the utilisation of existing resources, we have developed a Python and PostgreSQL application that is able to translate the existing information into a standardised database with a very well-defined data structure (https://github.com/bonacci-johannes/meda). Specifically, we inherit the individual centric view fundamental to medical science and attach additional information as separated tables that can be brought together to analyse various questions. These tables separate cross-sectional and longitudinal data and are grouped based on their clinical relevance. The typical database structure we have utilised is provided in Figure 1A and highlights the components that are required to be configured within our tool.

**FIGURE 1.**
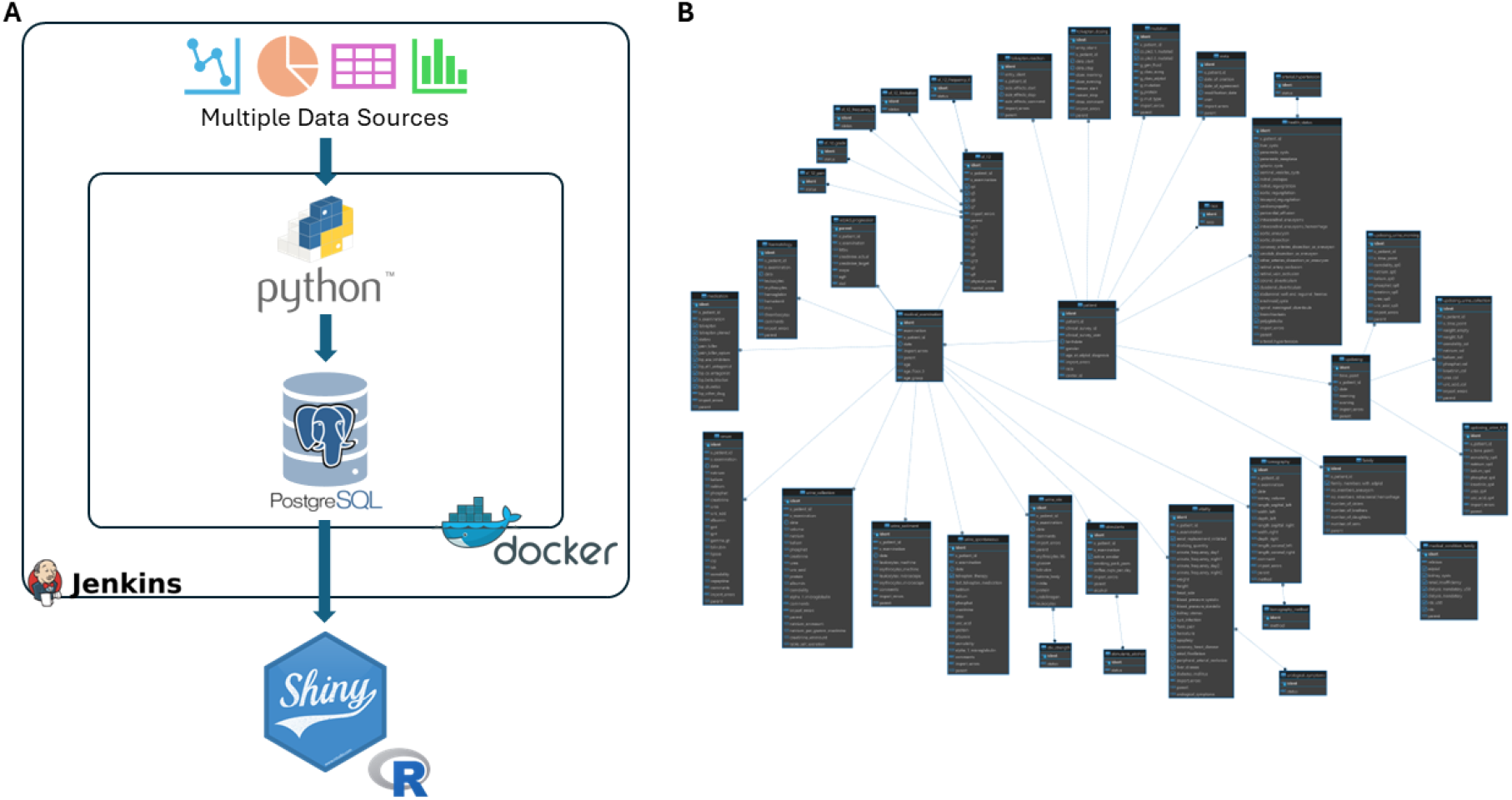
SCHEMATIC OVERVIEW OF MEDA. A) THE DEVELOPED PIPELINE AND CONTAINERS USED TO TRANSFORM CLINICAL DATA TO A REAL-TIME VISUALISATION PLATFORM. B) THE SCHEMATIC OVERVIEW OF THE POSTGRESQL DATABASE USED WITHIN MEDA.

To test this simple structure, we used a large patient cohort with chronic kidney disease available at the University Hospital Cologne and translated the currently utilized ClinicalSurveys.net (Vehreschild et al., 2010) database using our tool. ClinicalSurveys is a web-based tool to design and collect patient relevant data through a simple survey based tool. It allows collaboration across multiple sites in a secure manner and enables a systematic data collection. This cohort data contains numerous, meticulously collected patient information ranging from different levels of laboratory values, questionnaires, family history, tomography, or historic clinical information. All in all, over 2000 variables were represented within this ClinicalSurveys database structure. The design of this database followed a fully patient centric approach where longitudinal data was encoded as repeated variables within its single database. While this can be a reasonable approach to collect prospective data on individuals over a longer period of time, it can be quite error prone as, particularly, longitudinal data may be entered in the wrong section of the database skewing downstream analysis. In addition, the long list of variables can lead to an increase in human errors during entry where misplaced punctuation marks or swapping of variables may occur. Downstream analyses and visualisation may then be skewed by these data structures. Furthermore, quality assurance is more difficult to achieve since the large number of variables within a single database is challenging to evaluate for human individuals. Meda addresses some of these challenges in a semi-automated fashion. Most importantly, the tool automatically generates the database structure based on the configured data slots required. In essence, Meda follows a simple 5 steps approach:

### Step 1: Reading Source Data

The pipeline begins by reading raw source data in a flat structure, where each value occupies its own column.

### Step 2: Data Class Organization

The flat data is organized into nested data classes, which correspond to SQL-tables. When defining the data classes. At this point transformations or other computed variables can be generated through the provision of additional python functions.

### Step 3: Data Class Factory

The data class factory populates the nested data classes from the flat data structure.

### Step 4: DTO (Data-Transfer-Object) Factory

The DTO factory translates the nested data classes into DTOs that mirror the SQL structure.

### Step 5: DTO Registry

The DTO registry manages the DTO factory and database connection. It generates a DTO from a data class and writes it to the database.

In addition, Meda aims to generate subsets of manageable chunks of data, following clinically relevant chunks of information. Import classes are defined that ensure errors can be caught and that data is imported in the right format. In prospective studies, where data is continuously collected over long periods of time, we are therefore able to import data on a regular basis. Next, based on the result of the class import an additional error table is generated that allows users to visualise these import errors and address them accordingly. We found that the visual representation had a significant influence on the motivation of our staff to fix and remedy the shown errors. Lastly, we implemented a threshold-based value verification system which aims to identify values that we deem to be “not compatible with life” and which are sent back to the users for verification.

## Application

### Setting up Meda for semi-automated data entry

As in our example, ClinicalSurveys was used to house and collect the data from various collaborators, we used Dockers surrounding our database and Meda tool to simplify the setting up and destruction of the database. Simply put, the PostgreSQL database is fully refreshed upon each update that is being made. This ensured that only one true data source was available and reduced the need for verification of data entries within our database. To manage the automated setting up and destruction of the database Jenkins was used. The Jenkins Butler (*Jenkins*, 2011) monitors changes in the source code, scripts, and classes that are required and updates the database as soon as changes are observed. The total workflow using this approach takes less than 3 minutes and can therefore be performed as often as daily if new data are expected on a regular basis. The aforementioned classes need to be implemented to ensure that the right data is entered into the database. A simple Patient centric import of individual characteristics is shown in Code Section 1. The utilised Feature keyword here is an included separate class which provides the information on how to construct a dataclass from a data dictionary and how to import it into the SQL table. It enables the use of transformers, specifications of the input key, specifications of target table type (error, crossectional, or longitudinal), and the potential defaults to consider.

**Code Section 1.**
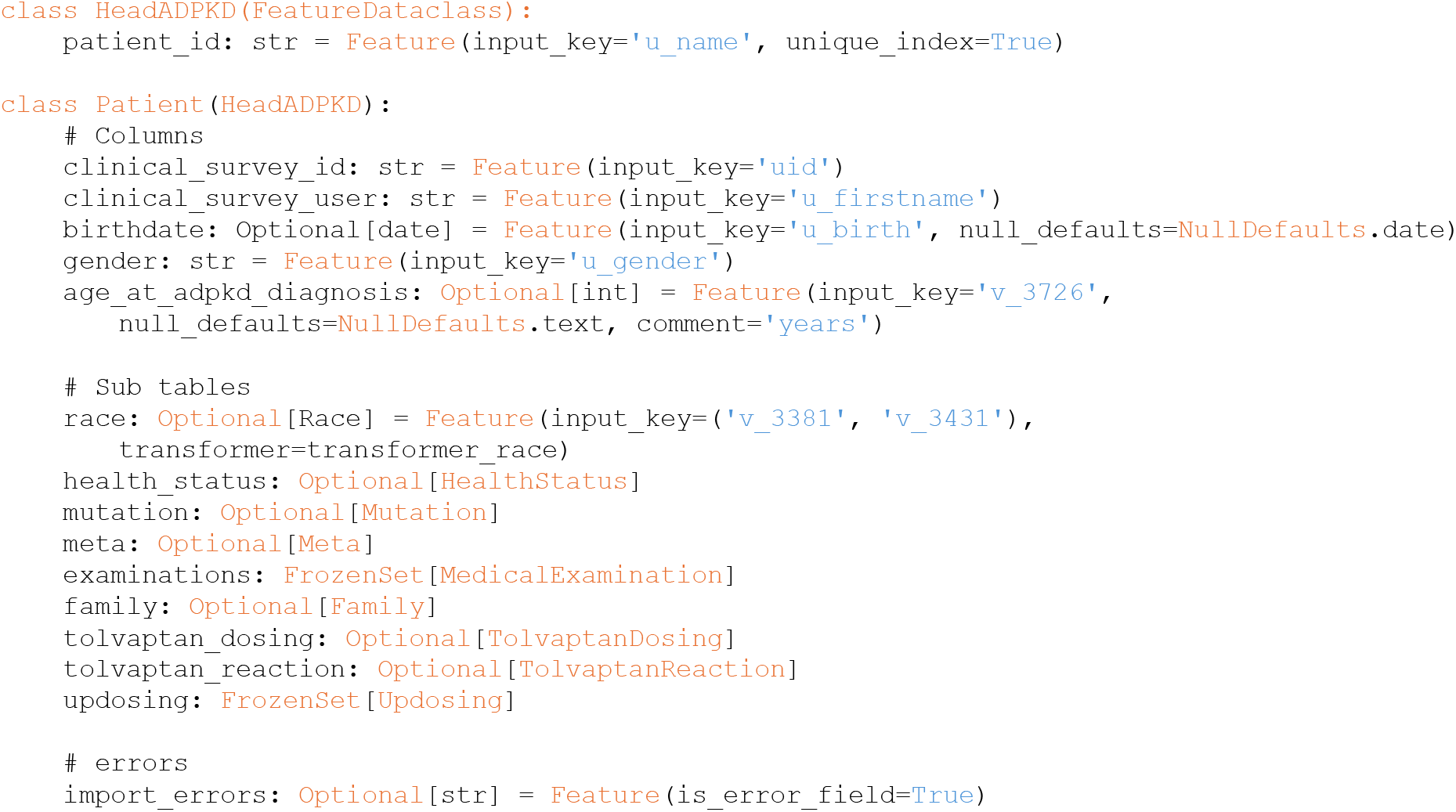
EXAMPLE CODE FOR EXTRACTING DATA INTO THE PROPOSED DATABASE SCHEMA.

### Automated evaluation and identification of missing and non-reliable data

During the data import, several additional steps are performed before adding the data to the database. First values are converted to a common reference unit. The unit conversion is a simple step but requires extensive configuration that covers all possible units. So far we have focused on the possible units within our CKD use case example and provide our conversion tables within the code. Code section 2 shows an example of such a configuration. This ensures that we do not need to store the unit information and that all data are converted to the relevant reference unit. Next, data are reviewed for known thresholds that are not compatible with life. Here a simple table (Figure 2), which can be adjusted by user dynamically through a web-based interface, is evaluated and any values exceeding these thresholds collected within their own separate table. The results are presented to the user who can then adjust, if necessary, the value within the original table used as input. This also applies to any missing data encountered during the data import.

**FIGURE 2.**
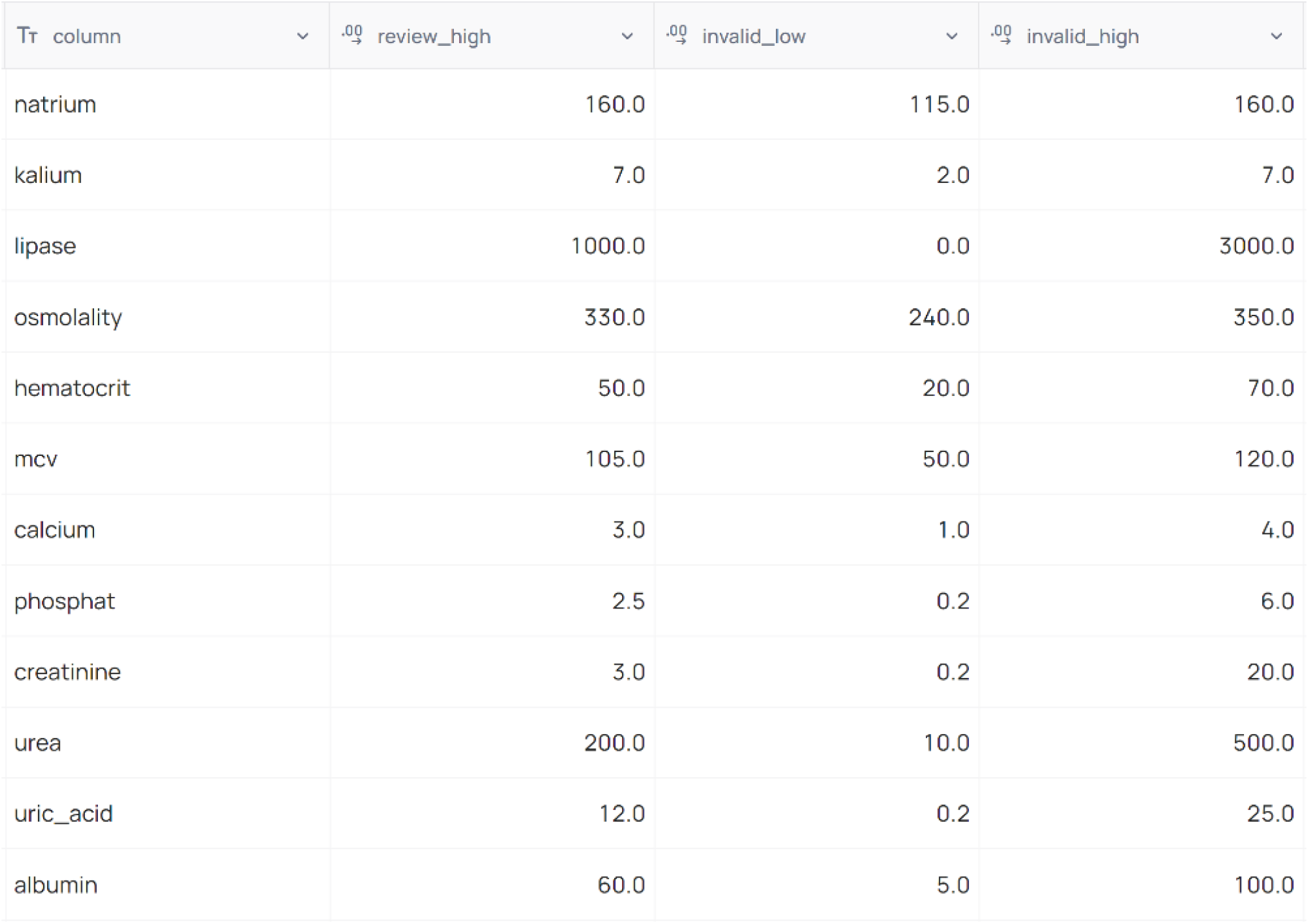
EXAMPLE THRESHOLDS TABLE USED DURING DATA IMPORT.

**FIGURE 3.**
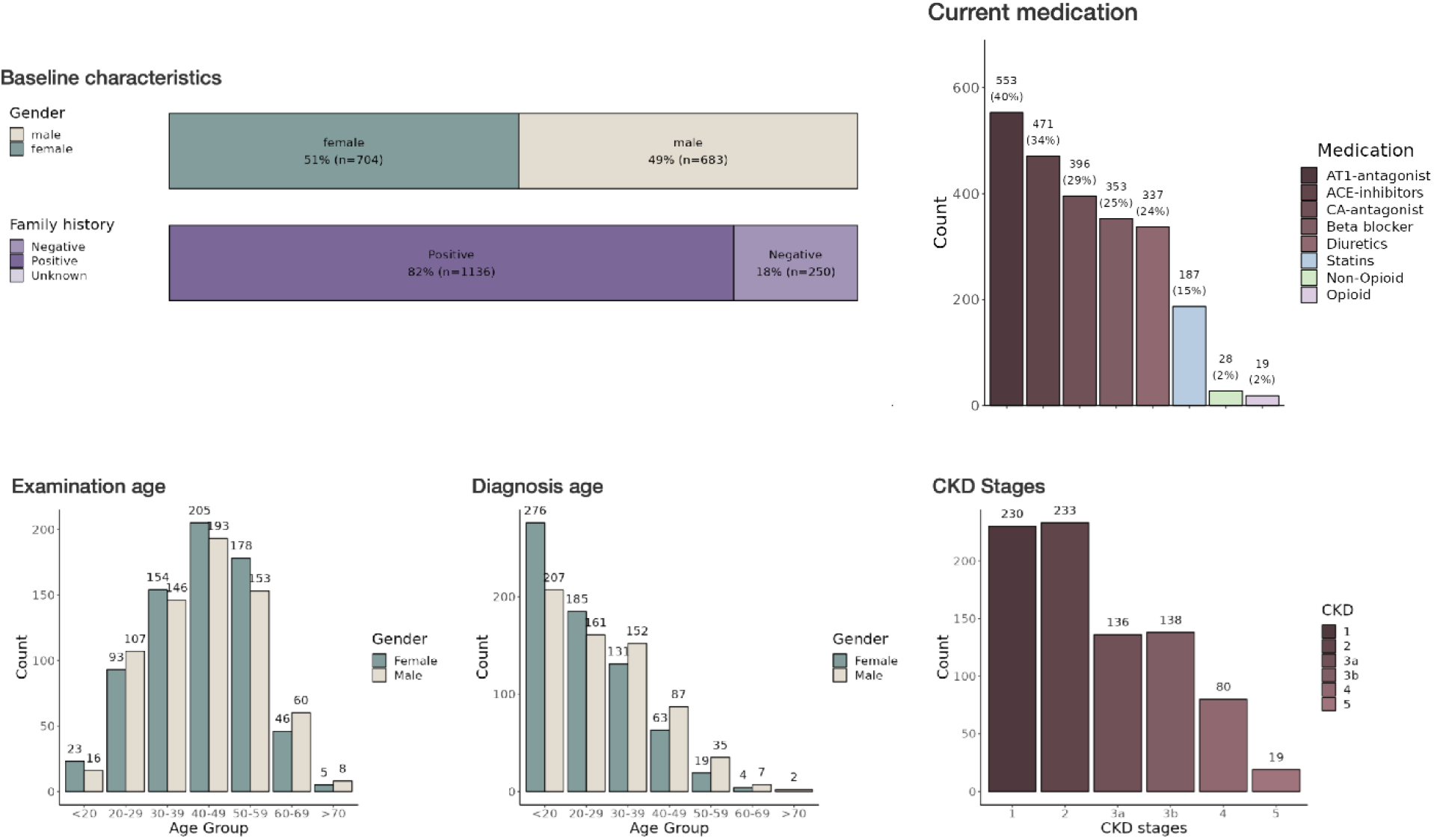
INTERACTIVE DASHBOARD OF THE ADPKD REGISTRY IN A SHINY APPLICATION – THIS FIGURE DISPLAYS PART OF THE INTERACTIVE DASHBOARD FROM THE SHINY APPLICATION USED TO VISUALIZE DATA FROM THE LARGE PATIENT COHORT WITH CHRONIC KIDNEY DISEASE. SEVERAL PAGES AND FILTER OPTIONS ARE AVAILABLE. THE DASHBOARD ITSELF IS SEGMENTED INTO TABS INCLUDING ‘PATIENT CHARACTERISTICS’, ‘DISEASE STAGE’, ‘MEDICATION’, AND OTHERS, ALLOWING FOR A COMPREHENSIVE OVERVIEW OF THE DATA CATEGORIES. THE ‘BASELINE CHARACTERISTICS’ SECTION PROVIDES HISTOGRAMS DETAILING DEMOGRAPHIC INFORMATION.

**Code Section 2.**
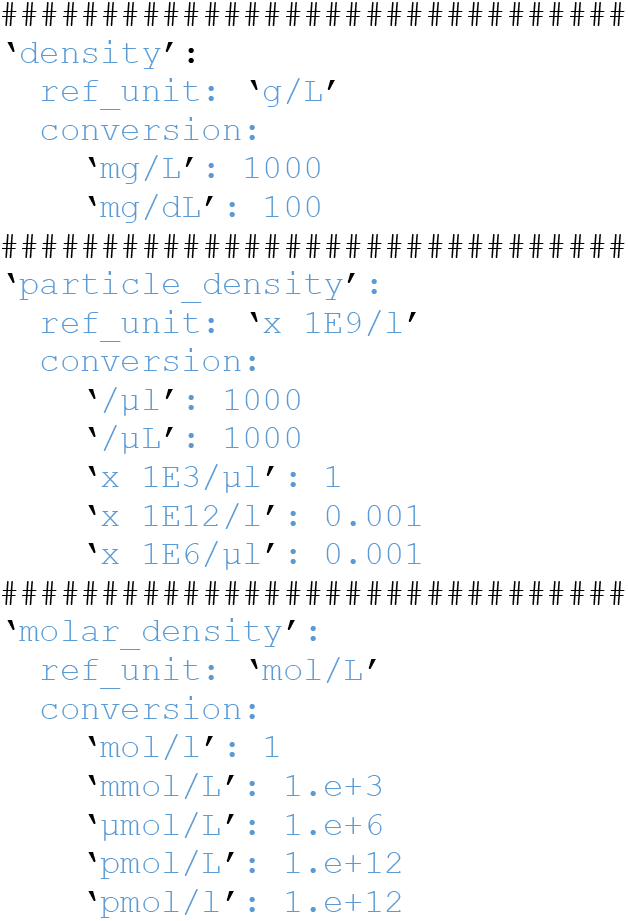
AUTOMATED CONVERSION OF UNITS DURING IMPORT AND PLAUSIBILITY CHECK.

This workflow can easily be integrated into daily clinical routines and allows for direct evaluation and visualization of the data. In addition, the near instant visual response to the fixing of missing or non-reliable data results in a significantly increased data quality. Furthermore, enabling auditing within PostgreSQL can provide a continuous log of changes that have been performed and ensure that data consistency is preserved.

### Visualisation and continuous evaluation of data provides new insights into patient health

The last step in our pipeline is the development of a visual representation of the data imported by Meda. Here we decided to develop an R (R Core Team, 2021) shiny application. While other types of frameworks exist to provide real-time views of such data, they are limited in their statistical analyses that can provide useful information in a clinical setting.

#### INTEGRATION AND FUNCTIONALITY OF SHINY APPLICATION IN R

The development of the Shiny application (Chang et al., 2024) represents a significant progression in the implementation of the ClinicalSurveys database. The Shiny framework in R facilitates the creation of dynamic web applications that offer the ability to visualize and analyze data in real time. Through the utilization of this technology, the application converts unprocessed clinical data into user-friendly, interactive dashboards and reports. As a result, in the future healthcare providers are provided with instantaneous access to patient information and trends. The Shiny application has been carefully designed to accommodate the particular requirements of healthcare professionals. The platform provides a collection of interactive tools that enable users to analyze demographic patient data along multiple axes, including time, disease advancement, and treatment results. At this degree of engagement, a more profound comprehension of patient health patterns is fostered, which empowers the development of individualized patient care plans and the discovery of effective treatment protocols.

#### CONTINUOUS EVALUATION FOR PROACTIVE HEALTHCARE

One of the most significant features of the Shiny application is its capability for continuous data evaluation. As the PostGreSQL database is refreshed with each update (once daily), the application automatically incorporates the latest data, ensuring that healthcare providers have access to the most current patient information. The insights garnered from the continuous evaluation of patient data have profound implications for both patient care and clinical research. For patient care, it enables a shift towards more proactive and personalized healthcare strategies, significantly improving patient outcomes. In the realm of research, the application provides a rich dataset for analyzing treatment efficacy, patient responses, and disease patterns, thereby contributing to the advancement of medical knowledge and the development of new treatment modalities.

## Discussion

The Meda pipeline was developed to bridge health registry data and real-time analysis of the available data. Our key approach was to develop a system where any type of clinical information could be imported, through the provision of simple configuration files, and where data could be displayed in near real-time to the user. Meda restructures and standardises such information and provides programmable access to this data. While we developed this in the context of clinical registries, its approach can be used for whole clinical databases that over the years have increased in complexity.

The choice of webfront was driven by the requirements within our statistical analyses. While there are a number of real-time visualisation frameworks available, such as Grafana, Metabase, or Tableau, they are not designed to handle clinical information and the underlying statistics within the biomedical domain. The shiny front, in combination with the many R packages available, allows us to generate and display any type of statistical analysis based on the data. Shiny, and therefore R, bring additional obstacles into this development as R is generally slow in utilising database queries, has a complex memory management, and can be inefficient in the use of data structures. To address these shortcomings we have opted to preprocess the database data every morning, and on demand, which generates the objects required for visualisation and statistical analysis and are loaded through serialized R object storage. This results in a much faster visualisation but limits the real-time application of our approach. Given that our registry data does not change on a daily basis and that data entry can be delayed based on clinical workload we struck a balance between functionality and overall speed in our approach. Further development of existing, faster, frameworks for visualisation would remedy this.

The growing need in clinical science to analyse and represent larger and more complex cohorts, with both retrospective and prospective data collection, has proved to be challenging due to the heterogeneity in collection systems, the lack of standardisation across healthcare institutions, and differences in ethical considerations. Our tool aims to address a number of these issues by enabling the integration and near real-time representation of data. By interfacing directly with a hospitals clinical data repository our tool could show important statistics and analyses in near real-time to clinical staff, ensuring an efficient and effective oversight of data entry in various settings. While raw data is the preferred data-type, the tool would also be able to collect already computed statistics and integrate data from multiple institutions to visualize the state of healthcare institutions over a larger geographical area while not exceeding the ethical considerations of each institution.

Overall we have established a tool that addresses the current scientific and clinical challenges in working with larger cohorts and provides a standardized structure for use within data science groups. We hope to enable a faster and simpler pipeline for clinical questions from data to results and drive the knowledge generation within medicine.

## Data Availability

All data produced in the present study are available upon reasonable request to the authors

## Notes

### Competing Interest Statement

The authors have declared no competing interest.

### Funding Statement

This study was funded by the Funded by the Deutsche Forschungsgemeinschaft (DFG, German Research Foundation) under Germany's Excellence Strategy, CECAD, EXC 2030, 390661388 and the Joerg Bernards Stiftung.

### Author Declarations

The German ADPKD registry (AD(H)PKD, NCT02497521), a prospective multicenter observational study that documents clinical and laboratory parameters as well as imaging data of study participants on an annual basis, was used to select the study population. The AD(H)PKD study enrolls adult patients (≥18 years) with ADPKD presenting for tolvaptan therapy evaluation. The study was approved by the local ethics committee of the University of Cologne and conducted in accordance with the Declaration of Helsinki and the International Conference on Harmonization's guidelines for good clinical practice.

